# Early individual and family predictors of weight trajectories from early childhood to adolescence: results from the Millennium Cohort Study

**DOI:** 10.1101/2020.02.26.20027409

**Authors:** Constança Soares dos Santos, João Picoito, Carla Nunes, Isabel Loureiro

## Abstract

**Background:** Early infancy and childhood are critical periods in the establishment of lifelong weight trajectories. Parents and early family environment have a strong effect on children’s health behaviors that track into adolescence, influencing lifelong risk of obesity.

**Objective:** We aimed to identify developmental trajectories of body mass index (BMI) from early childhood to adolescence and to assess their early individual and family predictors.

**Methods:** This was a secondary analysis of the Millennium Cohort Study and included 17,166 children. Weight trajectories were estimated using growth mixture modeling based on age- and gender-specific BMI Z-scores, followed by a bias-adjusted regression analysis.

**Results:** We found four BMI trajectories: Weight Loss (69%), Early Weight Gain (24%), Early Obesity (3.7%), and Late Weight Gain (3.3%). Weight trajectories were mainly settled by early adolescence. Lack of sleep and eating routines, low emotional self-regulation, child-parent conflict, and low child-parent closeness in early childhood were significantly associated with unhealthy weight trajectories, alongside poverty, low maternal education, maternal obesity, and prematurity.

**Conclusions:** Unhealthy BMI trajectories were defined in early and middle-childhood, and disproportionally affected children from disadvantaged families. This study further points out that household routines, self-regulation, and child-parent relationships are possible areas for family-based obesity prevention interventions.

## Introduction

Obesity is a major Public Health issue worldwide. Over the last decades, obesity has been increasing at an alarming rate and appearing at progressively younger ages (1). In fact, between 1975 and 2016, among children and adolescents aged 5–19 years, the global prevalence of overweight has increased from 4% to 18% in girls and 19% in boys, and the global prevalence of obesity has risen from 0.7% to 5.6% in girls and from 0.9% to 7.8% in boys, summing up to 340 million children and adolescents (1). Besides, 41 million children under 5 years of age were estimated to suffer from overweight or obesity in 2016 (2).

In the short term, children with overweight and obesity not only have a higher risk of hypertension, diabetes, and sleep problems, they also have a higher risk of psychological distress such as negative body image, low self-esteem, depression, and peer problems (3,4). Furthermore, unhealthy weight tends to persist into adolescence and adulthood, increasing the lifelong risk of these non-communicable diseases (5).

Early infancy and childhood appear to be critical periods in the establishment of lifelong weight trajectories (6–8). Therefore, it is important to understand these trajectories and their early determinants in order to inform effective public health interventions.

From an ecological perspective, family plays a major role in every aspect of a child’s health and development, especially during infancy and early childhood (9). Both observational and experimental studies support the persistent effect of early family environment on health behaviors and weight status, highlighting the central role of parents in childhood obesity (10). Several studies suggest that general parenting, parenting styles and practices, and parent-child relationships can shape early eating, exercise, sleep, and screen use habits that track into adolescence (11,12). However, its influence on weight status is still debated.

An empirical way to investigate body mass index (BMI) trajectories over time is by using Growth Mixture Modeling (GMM), an extension of Growth Curve Modeling (13). GMM is a person-oriented approach that assumes that individuals do not belong to a single homogenous population but rather to distinct unobserved subpopulations with different developmental trajectories (14). GMM focuses on longitudinal change within each subpopulation and allows the classification of individuals into latent classes based on their growth trajectories.

Investigating BMI trajectories of over time requires large samples and accurate anthropometric measurements. Although several studies explored weight trajectories in childhood, some studies used categorical measures of overweight and obesity, which leads to classification bias, and others used “crude” BMI values, which do not account for growth (13). Since normal growth results in an expected increase in BMI, age- and gender-specific BMI standard deviation scores (Z-scores) are often considered the gold standard for the analysis of anthropometric data at an epidemiological level (15). In fact, the Z-score has a linear scale that allows comparison between age groups and gender. Furthermore, it can be analyzed by summary statistics: the mean Z-score reflects the nutritional status of the entire population, and the standard deviation (SD) of the Z-score reflects the quality and accuracy of the data (15). On the other hand, most of these studies explored a small subset of covariates such as gender, ethnicity, and socioeconomic status, leaving out important factors like birthweight and breastfeeding duration that are well known to influence weight status, and even fewer studies have included early family environment, general parenting, and parenting practices.

Thus, the main objectives of our study were 1) to identify distinct subpopulations with different developmental trajectories of BMI in a general prospective cohort of British children, the Millennium Cohort Study; and 2) to examine the association between these trajectories and individual and family factors, focusing on those of early childhood.

In the literature, the most commonly found trajectory is the stable normative trajectory comprising the larger portion of the sample (13). While most studies report a decreasing BMI trajectory and an increasing BMI trajectory, other studies indicate an additional stable high trajectory (13).

Therefore, we hypothesize 1) the existence of different weight trajectories, including a stable normative trajectory and a persistent high trajectory; 2) that individual and family factors influence the risk of belonging to a group trajectory; and 3) that an adverse early family context increases the risk of following an unhealthy weight trajectory.

## Methods

### Case Study

Data were drawn from the Millennium Cohort Study (MCS), a cohort study that follows children born between September 2000 and January 2002, and living and growing up in England, Scotland, Wales, and Northern Ireland. It also provides information about family circumstances and the broader socioeconomic context. MCS was designed to overrepresent specific subgroups of the population, namely children living in disadvantaged areas and those who are ethnic minorities. The sample is clustered by electoral wards stratified by country, ethnicity, and Child Poverty Index (16).

The study began with an original sample of 18,552 families, and at Sweep 2 (2003–04), it recruited 691 “new families” who were eligible but were missed at Sweep 1. Therefore, the total number of families ever interviewed comprises 19,243 families (19,517 children). Children were around 9 months at Sweep 1 and about 3, 5, 7, 11, and 14 years old at the subsequent sweeps. The study protocol meets the ethical requirements of the Helsinki Declaration, and it was approved by the Northern and Yorkshire Research Ethics Committee (07/MRE03/32). Informed consent was obtained from parents or legal guardian before participation. Further information about the study design can be found in (16).

To the present study, singletons and the first-born child of the families with twins and triplets with valid data on BMI in at least one of Sweeps 2–6 were included, comprising 17,166 children.

### Measures

#### Anthropometric measures

Weight and height were measured by trained interviewers using standardized instruments (Tanita HD-305 scales, Tanita UK Ltd; and portable stadiometers, Leicester Height Measure, Seca UK), with children wearing neither shoes nor outdoor clothes. Weight and height were used to calculate BMI (kg/m^2^).

We calculated age- and gender-specific BMI z-scores (BMIz) using World Health Organization (WHO) Anthro (17) and Anthro Plus (18) software, having as reference the WHO Multicenter Growth Reference Study population. Observations with extreme values (below −5 SD or above

+5 SD) were considered outliers and excluded(15). Four individuals presented BMIz values only in the first two sweeps, with borderline BMIz at age 3 (around −4 SD) corresponding to extreme thinness, and at age 5 in the overweight range (around +1.5 SD). Since there were no subsequent values to validate these observations, they were considered highly implausible and excluded.

#### Covariates

Predictors of nutritional status were selected based on previous research, according to Davidson and Birch’s ecological model of overweight (9). A conceptual framework of early life predictors of Overweight and Obesity is presented in Figure 1. Further details on how covariates were categorized and coded are presented in Table S1.

**Figure 1.**
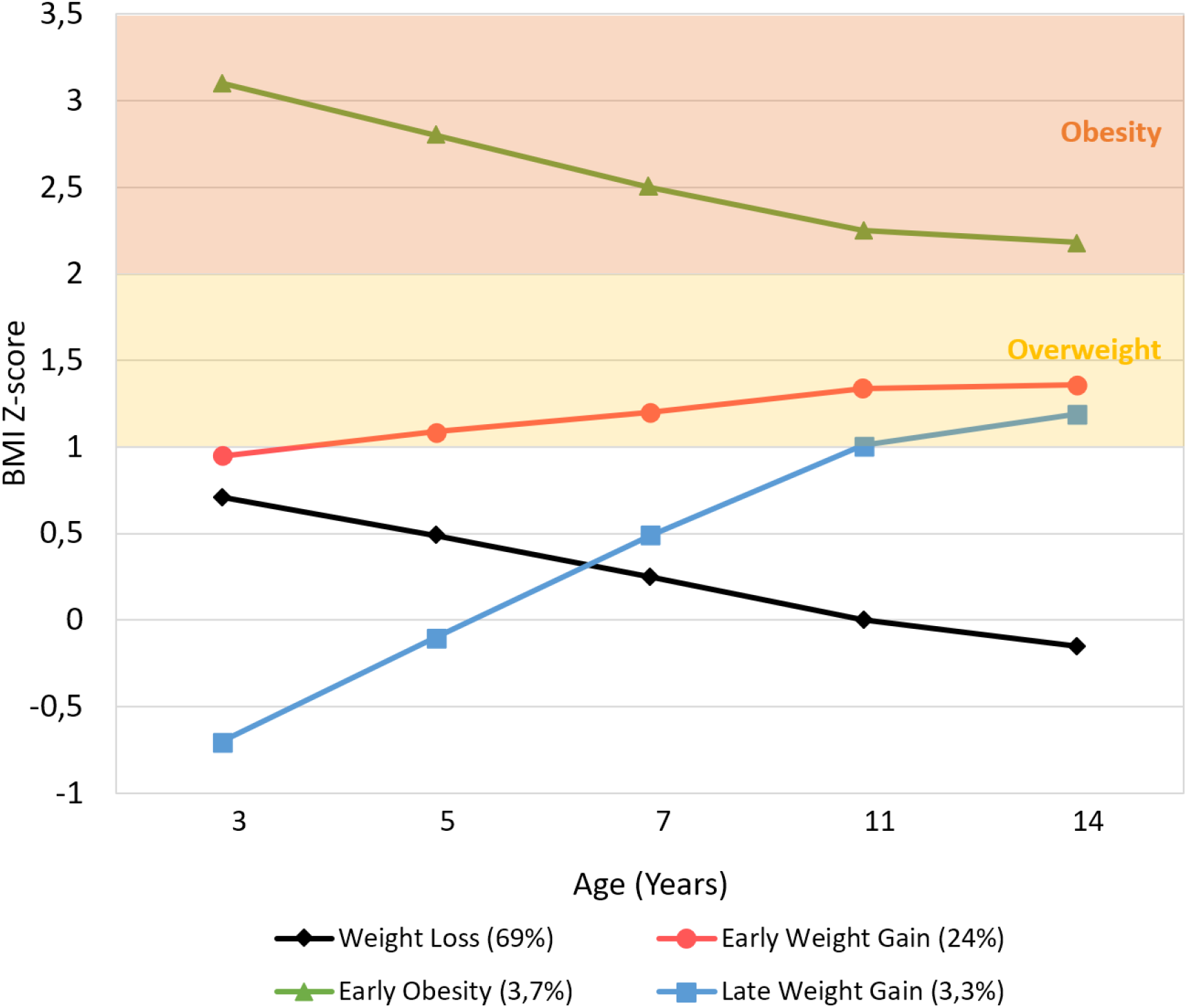
BMI z-score trajectories.

#### Sociodemographic and economic covariates

were gender, ethnicity, family structure, family poverty, maternal age at birth of cohort member, maternal nutritional status, and maternal education measured at 9 months.

#### Perinatal and early infancy covariates

<u>Gestational age</u> (GA) was categorized as “extreme to moderate prematurity,” “late prematurity,” and “term,” according to the WHO/UNICEF definition (19).

<u>Birthweight</u> was categorized as “low birthweight,” “normal birthweight,” and “high birthweight,” according to the WHO definition (20).

<u>Birthweight</u> centiles for age and gender were calculated using Intergrowth21 software, and individuals were further classified in “Appropriate for GA,” “Small for GA,” and “Big for GA” (21).

<u>Breastfeeding duration</u> was assessed in Sweeps 1 and 2; it was re-categorized as in (22), with two more categories (“6 to 12 months” and “more than 12 months”), according to its distribution.

<u>Introduction to solid food</u> was classified as “early introduction” if it occurred before 4 months, and “late introduction” if it occurred after 6 months, according to European Society for Pediatric Gastroenterology Hepatology and Nutrition recommendations (23).

#### Child temperament and self-regulation

<u>Child temperament</u> was assessed at 9 months by 14 items from the Carey Infant Temperament Scale, capturing three dimensions: mood, adaptability/approach-withdrawal, and regularity. We created total scores for each dimension by summing the individual responses, as in (24). High scores on the first two dimensions indicated distress and withdrawal (α = .546 and .677, respectively), and high scores on the last dimension indicated regularity (α = .713).

<u>Child self-regulation</u> was assessed at 9 months by 10 items from the Child Social Behavior Questionnaire, with two dimensions: cognitive (α = .573) and emotional self-regulation (α = .632). Total scores for each dimension were calculated as the average of valid responses (25), so that higher scores in the first dimension indicated higher cognitive self-regulation, and higher scores in the second indicated emotional dysregulation.

#### Household routines, parenting beliefs, and parenting activities

<u>Household routines</u> were assessed at age 3, comprising sleep and feeding routines. A total sum score was created, with higher scores indicating consistent routines (α = .541).

<u>Parenting beliefs</u> were assessed at 9 months comprising 3 items derived from the European Longitudinal Study of Pregnancy and Childhood about the importance of stimulating, talking, and cuddling to a baby’s development. We summed the responses (26), so that higher scores indicated more positive parenting beliefs (α = .730).

<u>Parenting activities</u> were assessed at age 3 and comprised 5 items (reading, teaching the alphabet, teaching counting, teaching songs/rhymes, drawing). A total sum score was generated, so that higher scores indicated higher involvement in these activities (α = .610).

#### Discipline practices and child-parent relationship

<u>Discipline practices</u> were assessed at age 3 by 6 items from Straus’s Conflict Tactics Scale, measuring how often the mother used punishing (smack, shout, “tell off”) or withdrawal tactics (ignore, take away treats, send to bedroom/naughty chair) when the child misbehaved. We created two scores: a harsh parenting score (α = .657) by summing the responses in the punishing items, and a positive parenting score (α = .556) by summing the responses in the withdrawal items (27).

<u>Parent-child relationship</u> was assessed at age 3 by Pianta Short Form, a 15-item self-administered scale based on attachment theory. This scale comprised two scores: closeness (α = .657) and conflict (α = .787).

### Statistical analysis

Statistical analysis was performed using IBM Statistical Package for the Social Sciences, version 24.0 (SPSS Inc., Chicago, IL) and Mplus, version 8.3 (Muthén & Muthén 2017). Statistical significance was set to *p* < 0.05.

#### Estimation of BMI trajectories

Longitudinal BMI trajectories were analysed with GMM. GMM classifies children into latent classes based on their longitudinal change, so that individuals with similar BMI trajectories are assigned to the same class, and individuals in different classes follow significantly different BMI trajectories (14).

We tested multiple GMM models with different specifications before choosing the final model. First, we performed single-group models to identify the pattern (intercept only, linear, quadratic, cubic) that represented change over time the best. We also performed a latent basis model, where the pattern of change is not predefined but driven by the data, and loadings for the *slope* factor are estimated to represent the proportion of the total amount of growth that has occurred up to that point. Next, we performed GMM with *intercept* and *slope* variances fixed at zero for each class, a subtype of GMM called Latent Class Growth Analysis (LCGA), which assumes that all individual growth trajectories within a class are homogeneous (14). Then, we performed several GMMs with different growth factor variance specifications: first, with equal variances (homoscedastic model), and then freely estimated (heteroscedastic model). To determine the optimal number of classes, we started with a one-class solution and progressively increased the number of classes.

#### Model selection

The best model was chosen considering information criteria, theoretical justification, and interpretability (28,29). First, we looked at two model fit indices: Schwarz’s Bayesian Information Criterion (BIC) and the Bootstrapped Likelihood Ratio Test (BLRT) (28). BIC considers the likelihood of a model as well as the number of estimated parameters, with lower values of BIC indicating better fitting. BLRT compares a model with k classes to a model with k-1 classes, providing a *p-*value. We then looked at entropy, a measure of classification quality and separation between classes. Higher values of entropy (near 1) indicate more confidence in the classification (30). Although values above 0.8 are considered good, there is no consensual definition of what constitutes low entropy (30). Finally, we considered the interpretability of the models and their practical meaningfulness, rejecting models with clinically uninterpretable classes and with classes representing less than 1% of the total sample.

#### Association between BMI trajectories and early individual and family factors

The association between covariates and class membership was calculated based on crude and adjusted odds ratio (OR) using a *bias-adjusted three-step approach*, which takes into account the classification error in class assignment (31).

#### Missing values

Missingness resulted from item non-response and attrition. Attrition is mainly related to sociodemographic characteristics, so the MCS study was designed to account for this bias and still provide representative information.

Missing values on BMI variables were handled with full information maximum likelihood (FIML) estimation under missing data theory. Considered the gold standard for handling missing data in latent variable indicators, FIML uses all available data points and is robust to non-normal distribution. (32)

Missing values on covariates were handled using multiple imputations carried out in Mplus. We used Bayesian estimation to create 25 imputed datasets, using the Markov chain Monte Carlo algorithm, and convergence criterion was set to 0.05. We included all covariates and also BMIz variables under the missing at random assumption, accounting for the complex sample design (32). Imputed values compare reasonably to those observed.

## Results

### Body Mass Index

Table 1 shows the summary statistics of BMIz across all sweeps and the nutritional status according to WHO BMIz cut-offs.

**Table 1.**
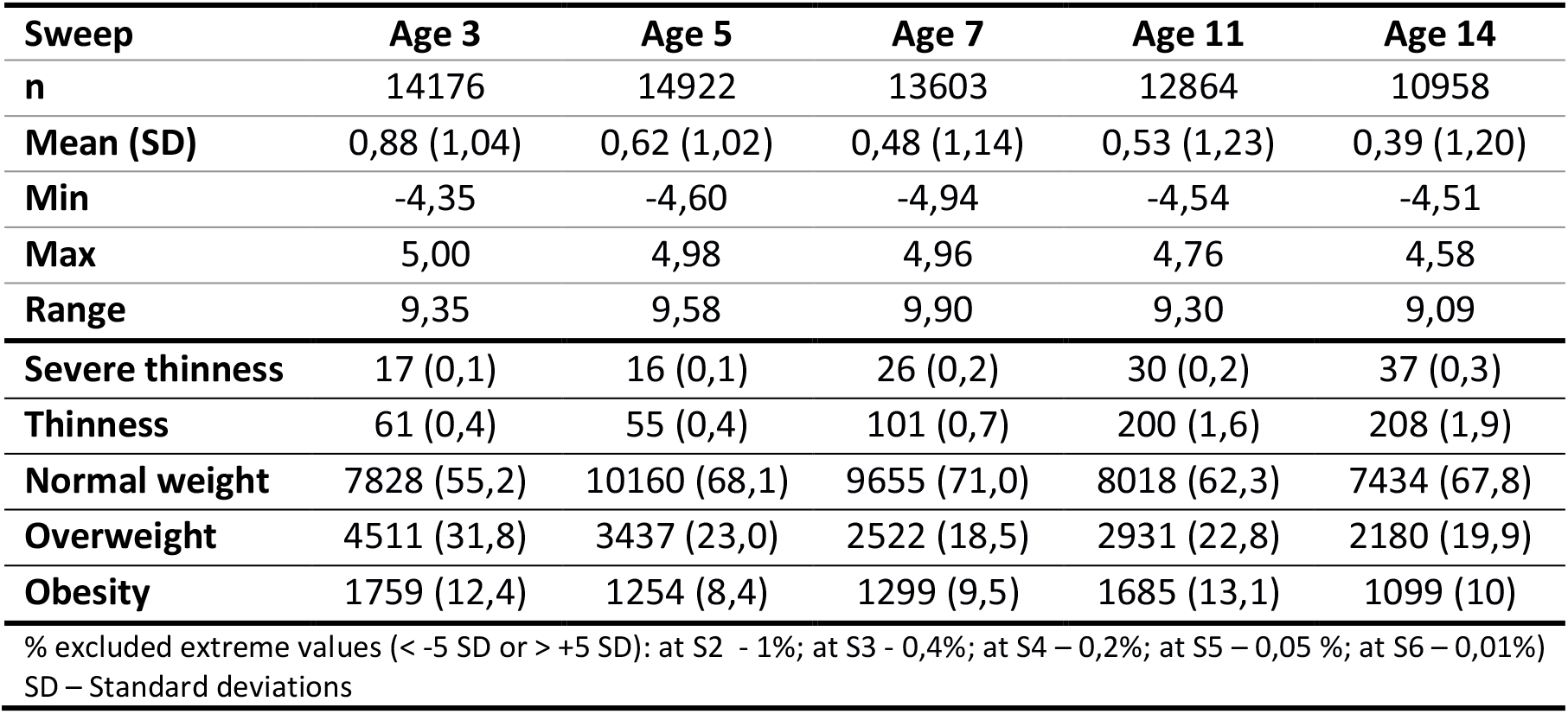
BMI z-score summary and descriptive statistics according to WHO cut-offs.

|  |  |  |  |  |  |
| --- | --- | --- | --- | --- | --- |
| **Sweep** | **Age 3** | **Age 5** | **Age 7** | **Age 11** | **Age 14** |
| **n** | 14176 | 14922 | 13603 | 12864 | 10958 |
| **Mean (SD)** | 0,88 (1,04) | 0,62 (1,02) | 0,48 (1,14) | 0,53 (1,23) | 0,39 (1,20) |
| **Min** | -4,35 | -4,60 | -4,94 | -4,54 | -4,51 |
| **Max** | 5,00 | 4,98 | 4,96 | 4,76 | 4,58 |
| **Range** | 9,35 | 9,58 | 9,90 | 9,30 | 9,09 |
| **Severe thinness** | 17 (0,1) | 16 (0,1) | 26 (0,2) | 30 (0,2) | 37 (0,3) |
| **Thinness** | 61 (0,4) | 55 (0,4) | 101 (0,7) | 200 (1,6) | 208 (1,9) |
| **Normal weight** | 7828 (55,2) | 10160 (68,1) | 9655 (71,0) | 8018 (62,3) | 7434 (67,8) |
| **Overweight** | 4511 (31,8) | 3437 (23,0) | 2522 (18,5) | 2931 (22,8) | 2180 (19,9) |
| **Obesity** | 1759 (12,4) | 1254 (8,4) | 1299 (9,5) | 1685 (13,1) | 1099 (10) |
| % excluded extreme values (< -5 SD or > +5 SD): at S2 - 1%; at S3 - 0,4%; at S4 - 0,2%; at S5 - 0,05 %; at S6 - 0,01%)<br>SD - Standard deviations |  |  |  |  |  |

Comparing our results to those of the WHO Multicenter Growth Reference Study, at age 3 our study sample showed a slightly higher mean BMIz, implying an upward shift of our sample distribution, with a progressive decrease in subsequent sweeps. The BMIz SD superior to 1 indicated a slightly more dispersed distribution, but in all sweeps, it was below 1.3, suggesting good quality of the data. The overall prevalence of overweight and obesity decreased from 44.2% at age 3 to 29.9% at age 14.

### Covariates

Sociodemographic characteristics of the analytical sample are presented in Table 2, and psychosocial covariates are presented in Table S2. Compared with children in the analytic sample, excluded individuals (who had no BMIzs in any of the sweeps) were more likely to be male, to be from ethnic minorities, to come from economically disadvantaged families and from monoparental families. Mothers of excluded children were younger and less educated. Complete bias analysis is available in Table S3.

**Table 2.** Characteristics of the analytical sample (n= 17166)

| **Table 2 – Characteristics of the analytical sample (n= 17166)** |  |  |  |  |
| --- | --- | --- | --- | --- |
|  |  | n | n (%) | Missing (%) |
| **Gender** |  | 17166 |  | 0 |
|  | Male |  | 8775 (51,1) |  |
|  | Female |  | 8391 (48,9) |  |
| **Ethnicity** |  | 17061 |  | 0,6 |
|  | White |  | 1405 (82,3) |  |
|  | Other |  | 3016 (17,7) |  |
| **Gestational age** |  | 16277 |  | 5,2 |
|  | Extreme to moderate Prematurity |  | 319 (2,0) |  |
|  | Late Prematurity |  | 892 (5,5) |  |
|  | Term |  | 15066 (92,6) |  |
| **Birthweight** |  | 17039 |  | 0,7 |
|  | Low |  | 1213 (7,1) |  |
|  | Normal |  | 14010 (82,2) |  |
|  | High |  | 1816 (10,7) |  |
| **Birthweight for gestational age** |  | 15959 |  | 7 |
|  | Small |  | 1237 (7,8) |  |
|  | Appropriate |  | 11884 (74,5) |  |
|  | Large |  | 2838 (17,8) |  |
| **Breastfeeding duration** |  | 17075 |  | 0,5 |
|  | Never |  | 5706 (33,4) |  |
|  | <=2 months |  | 4951 (29) |  |
|  | 2 to 4 months |  | 2067 (12,1) |  |
|  | 4 to 6 months |  | 1270 (7,4) |  |
|  | 6 to 12 months |  | 1668 (9,8) |  |
|  | >=12 months |  | 1413 (8,3) |  |
| **Age at complementary feeding** |  | 16468 |  | 4,1 |
|  | Before 4 months |  | 5146 (31,2) |  |
|  | Between 4 and 6 months |  | 10836 (65,8) |  |
|  | After 6 months |  | 486 (3) |  |
| **Family structure (Sweep 1)** |  | 16486 |  | 4 |
|  | Two parents/ carers |  | 13766 (83,5) |  |
|  | Single parenthood (One parent/ carer) |  | 2720 (16,5) |  |
| **OECD Median Poverty Indicator (Sweep 1)** |  | 16429 |  | 4,3 |
|  | Above 60% median |  | 10528 (64,1) |  |
|  | Below 60% median |  | 5901 (35,9) |  |
| **Mother age at cohort member's birth** |  | 17101 |  | 0,4 |
|  | 12 to 19 years |  | 1476 (8,6) |  |
|  | 20 to 39 years |  | 15257 (89,2) |  |
|  | 40 plus |  | 368 (2,2) |  |
| **Mother nutritional status (Sweep 1)** |  | 14297 |  | 16,7 |
|  | Underweight |  | 1825 (12,8) |  |
|  | Normal weight |  | 6872 (48,1) |  |
|  | Overweight |  | 3721 (26) |  |
|  | Obesity |  | 1879 (13,1) |  |
| **Maternal education (Sweep 1)** |  | 16432 |  | 4,3 |
|  | Low (NVQ<3 / none/ overseas) |  | 11492 (69,9) |  |
|  | High (NVQ=>4) |  | 4940 (30,1) |  |
| **Data are given as n (%)** |  |  |  |  |
| **NVQ – National Vocational Qualifications** |  |  |  |  |

### BMI trajectory estimation

The single-group models that served as a basis for subsequent GMM are presented in Figure S2, showing that the quadratic and latent basis models appeared to better explain change in BMIz over time. Although the Quadratic GMM showed lower BICs, they provided uninterpretable trajectories; therefore, we decided to further analyze the latent basis GMM, presented in Table 3.

**Table 3.** Growth Mixture Models.

| Latent Basis Latent Class Growth Analysis<br>(growth factor variances fixed at 0) |  |  |  |  |  |
| --- | --- | --- | --- | --- | --- |
| No of classes | 1 | 2 | 3 | 4 | 5 |
| Log-likelihood | -101683.801 | -90860.327 | -85782.504 | -83534.897 |  |
| AIC | 203387.602 | 181746.654 | 171597.007 | 167107.794 |  |
| BIC | 203465.108 | 181847.412 | 171721.017 | 167255.056 |  |
| aBIC | 203433.329 | 181806.099 | 171670.170 | 167194.675 |  |
| Entropy | - | 0.74 | 0.78 | 0.77 |  |
| BLRT *p*-value | - | <0.001 | <0.001 | 1.0000 |  |
| Parameters | 10 | 13 | 16 | 19 |  |
| n | 17166 | 10352/ 6813 | 2895/ 4803/<br>9467 | 2484/ 1393/<br>5195/ 8093 |  |
| Latent Basis Homoscedastic Growth Mixture Model<br>(growth factor variances equal between classes) |  |  |  |  |  |
| No of classes | 1 | 2 | 3 | 4 | 5 |
| Log-likelihood | -79740.295 | -79489.756 | -79363.766 | **-79334.793** | -79306.347 |
| AIC | 159506.591 | 159011.511 | 158765.531 | **158713.586** | 158662.694 |
| BIC | 159607.349 | 159135.521 | 158912.793 | **158884.100** | 158856.460 |
| aBIC | 159566.035 | 159084.674 | 158852.413 | **158814.185** | 158777.011 |
| Entropy | - | 0.88 | 0.82 | **0.60** | 0.65 |
| BLRT *p*-value | - | <0.001 | <0.001 | **<0.001** | <0.001 |
| Parameters | 13 | 16 | 19 | **22** | 25 |
| n | 17166 | 16689 / 476 | 519 / 408 /<br>16238 | **12600/ 3657/**<br>**505 / 403** | 12728/ 3539/<br>510/ 357/ 31 |
| Latent Basis Heteroscedastic Growth Mixture Model<br>(growth factor variances variant between classes) |  |  |  |  |  |
| No of classes | 1 | 2 | 3 | 4 | 5 |
| Log-likelihood | -79740.295 | -79341.995 | -79272.012 | -79248.190 |  |
| AIC | 159506.591 | 158721.990 | 158594.023 | 158558.380 |  |
| BIC | 159607.349 | 158869.252 | 158787.789 | 158798.649 |  |
| aBIC | 159566.035 | 158808.871 | 158708.341 | 158700.133 |  |
| Entropy | - | 0.49 | 0.48 | 0.46 |  |
| BLRT *p*-value | - | 0.0000 | 1.0000 | 1.0000 |  |
| Parameters | 13 | 19 | 25 | 31 |  |
| n | 17166 | 14482/ 2683 | 13870/ 2943/<br>352 | 12981/ 2567/<br>915/ 702 |  |

BLRT = Bootstrapped Log-Likelihood Ratio Test
All Growth Mixture Models were estimated without covariates using MPlus, version 8.3 (ANALYSIS = MIXTURE; ALGORITHM = INTEGRATION). The number of random starts was increased to 2000 with 500 final stage optimizations (STARTS = 2000 500). To compute the BLRT, the number of random starts for the k class was increased to 200 with 40 final stage optimizations (LRTSTARTS = 0 0 200 40). Then, we repeated the analysis accounting for the complex sample design (ANALYSIS = MIXTURE COMPLEX).

The <u>LCGA</u> showed much higher BICs than the other models, and since LCGA is based on the assumption that there is no within-class variability, we did not explore these models further.

Regarding the <u>homoscedastic GMM</u>, the *2-class model* showed a big heterogeneous class (95.6%) and a small well-defined class (4.4%), corresponding to an Early Obesity trajectory class.

The *3-class model* provided classes that were very disproportional (one big class comprising 91.9% and the two remaining classes comprising 4.3% and 3.8%). Furthermore, the two smaller classes were interpretable and meaningful, corresponding to an Early Obesity trajectory (4.3%) and a Late Weight Gain trajectory (3.8%); however, the bigger class (91.9%) was still heterogeneous, showing 20% of overweight and obese children across all sweeps, not corresponding to a normative class that we could use as a reference.

The *4-class model* showed more proportional and meaningful classes; it provided an Early Obesity trajectory (3.7%) and a Late Weight Gain trajectory (3.3%) similar to the 3-class model, and Weight Loss (69.0%) and Early Weight Gain (24%) trajectories. The *5-class model* showed an additional class that represented only less than 1% of the sample.

Regarding the <u>heteroscedastic GMM models</u>, they showed lower BIC, with BLRT favoring the 2-class model. This model provided a decreasing trajectory (77.7%) and an increasing trajectory (22.3%). However, this model showed low entropy (0.49) and provided poorly defined classes, while the decreasing trajectory still comprised around 20% of overweight and obese children at Sweep 6.

Therefore, we considered the 4-Class Latent Basis Homoscedastic GMM the best model to explain the change in BMIz.

### BMI trajectory characterization

Figure 1 shows the BMIz trajectories from early childhood to adolescence. Latent growth factor means and variances are available in Table S4 for each class. The *intercept* and *slope* are inversely correlated (*r* = −0.257, *p* < 0.001), meaning that the higher the initial BMIz, the lower the growth. The time scores are 0, 0.326 (*p* < 0,001), 0.633 (*p* < 0,001), 0.924 (*p* < 0,001), and 1, meaning that individuals have reached 32.6% of the total change in BMIz at age 5, 63.3% at age 7, 92.4% at age 11, and 100% at age 14. Thus, the greatest amount of change occurs during early and middle-childhood, and there is little further change (7.6%) in BMIz in early adolescence.

Observing the BMI trajectories, two clearly distinct classes are seen: the Early Obesity and the Late Weight Gain. The Early Obesity class follows a clearly distinct trajectory across all sweeps, with a significantly higher mean BMIz at the starting point (mean intercept = 3.102, *p* < 0.001) than the other classes, that then decreases until the end of the study period (mean slope = −0.935, *p* < 0.001) but is still above the obesity cut-off. Although the Late Weight Gain class shows a lower starting point than the other classes (mean intercept = −0,716, *p* < 0.001), it then shows the greatest increase (mean slope = 1.889, *p* < 0.001) throughout early and middle-childhood, reaching the cut-off of overweight by age 11.

The other two bigger classes (Weight Loss and Early Weight Gain) have close starting points at age 3 (normal-high mean intercepts of 0.723 and 0.931, respectively; Wald test *p* = 0.06), but from then forward, they follow significantly opposite trajectories during childhood (Wald test *p* < 0.001): the Weight Loss Class steadily decreases (mean slope = −0.781), remaining in the normative BMIz range, while the Early Weight Gain increases (mean slope = 0.428) and plateaus in the overweight range.

The proportions of overweight and obese children in each class is represented in Figure 2.

**Figure 2.**
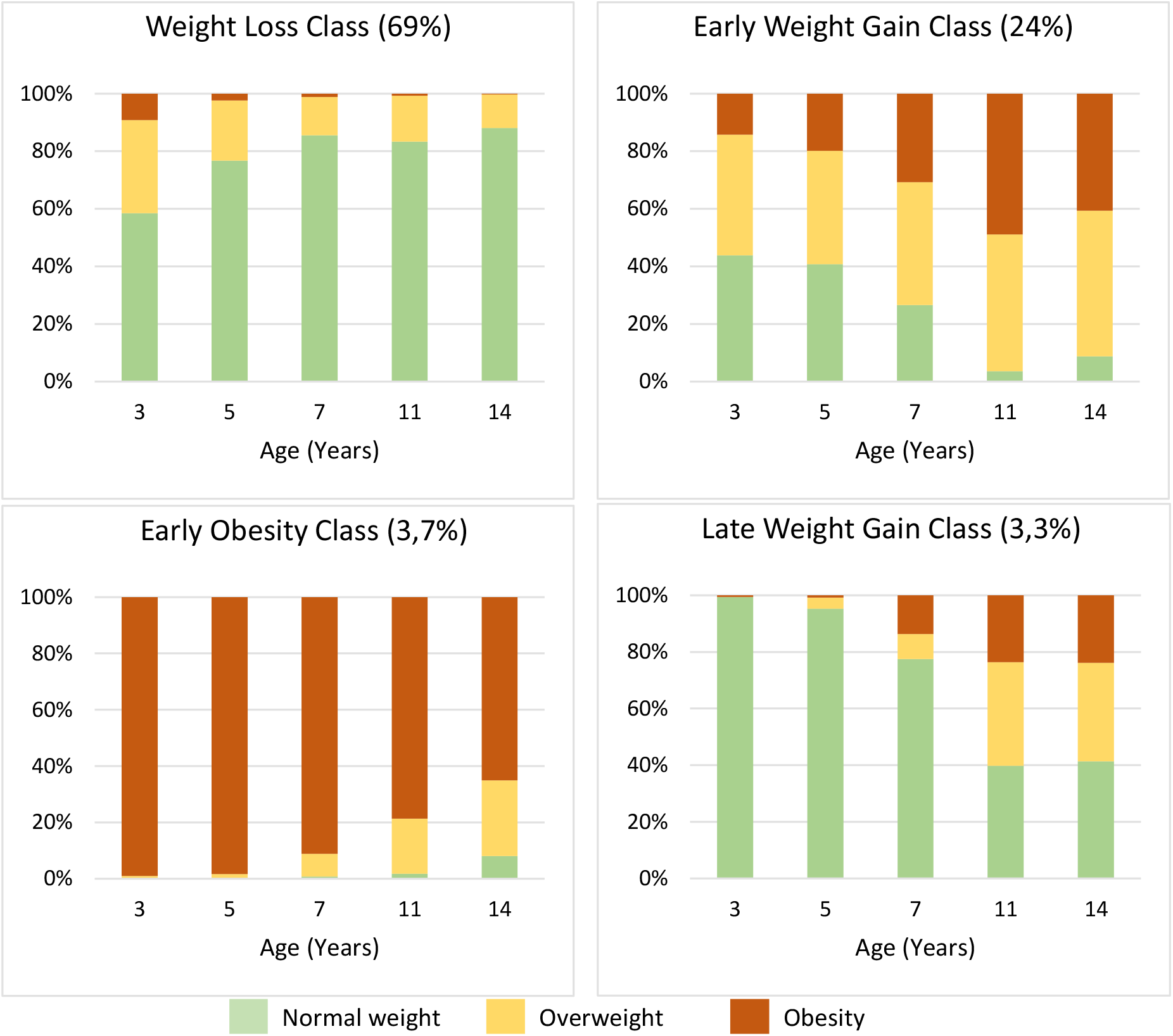
Class characterization – nutritional status.

### Association between BMI trajectories and individual and family covariates

The association between BMI trajectories and individual and family covariates are shown in Figure 3, and unadjusted and adjusted OR are available in Table S5.

**Figure 3.**
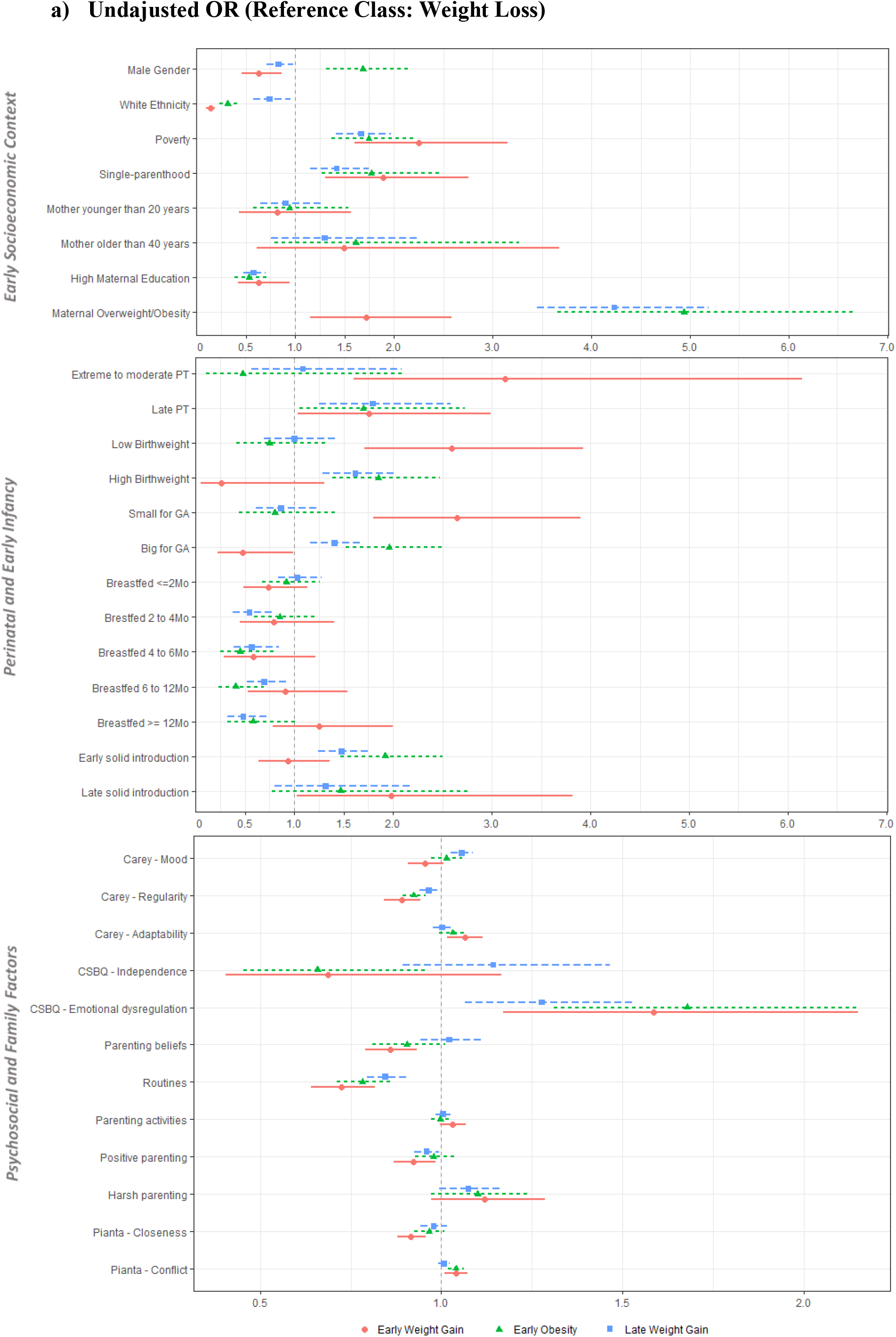
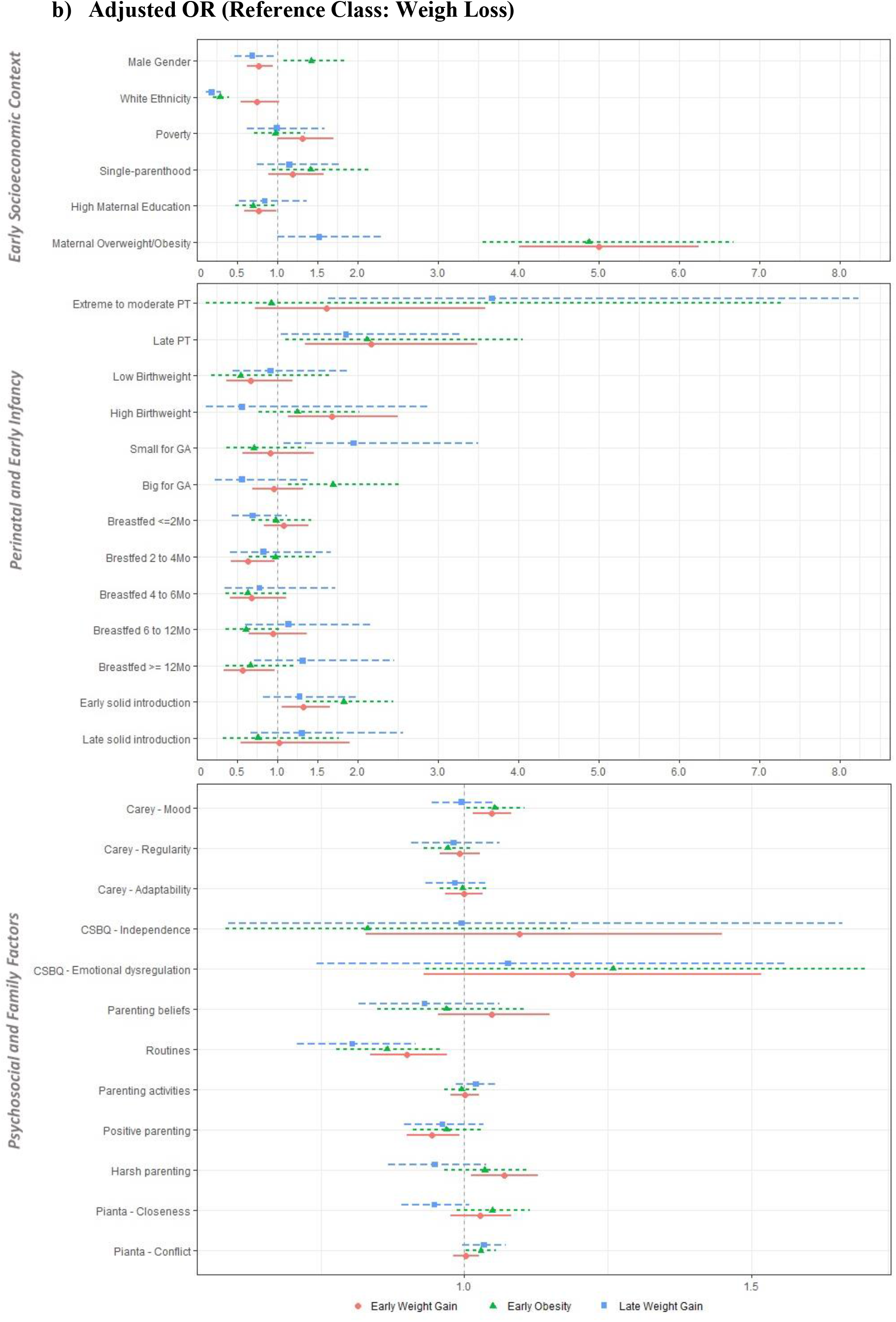
Early Life Predictors of BMI trajectories. CSBQ – Child Social Behavioural Questionaire; GA – Gestational Age; PT – prematurity

## Discussion

We used GMM to capture the developmental change in BMI from early childhood to adolescence. We found four weight trajectories: Weight Loss (69%), Early Weight Gain (24%), Early Obesity (3.7%), and Late Weight Gain (3.3%). Using data from the MCS, a recent study applying LCGA to raw BMI data from 3 to 11 years also found four trajectories: *Stable* (83.8%), *Moderate Increasing* (13.1%), *High Increasing* (2.5%), and *Decreasing* (0.6%) (33). In addition, a similar study using the same period and that applied GMM instead of LCGA to raw BMI values found four trajectories, but with a significantly different interpretation: *Low normal* (boys, 49%; girls, 42%), *Mid normal* (boys, 36%; girls, 38%), *Overweight* (boys, 12%; girls, 16%), and *Obesity* (boys, 2%; girls, 3%) (34). In the latter study, there were no increasing or decreasing trajectories, meaning that children did not change to different weight categories during the study period (no mobility from normal to overweight/obesity categories or vice-versa) (34). Based on a methodologically different approach, our study further builds on the contrasting results of these previous studies, applying a homoscedastic latent basis GMM on age- and gender-specific BMIz to better capture BMI variation with growth and expanding the analysis from age 3 to age 14.

### BMI trajectories are mainly settled by early adolescence

The greatest amount of change in BMIz occurred during early and middle-childhood, and there was little further change after age 11. In our study sample, by the time children enter school, they have reached one third of the total amount of change in their BMIz; further, 59.8% of the total amount of change occurs between ages 5 and 11, suggesting that early and middle-childhood are two different critical periods to intervene in weight trajectories. The importance of preschool years in weight gain has been established in other studies (6). In contrast, in one study using data from the Avon Longitudinal Study of Parents and Children (7) and in another in the USA (8), BMIzs steadily increased during childhood, with the greatest change occurring after school entry, suggesting that excess weight gain and obesity also develop in middle-childhood. In both studies, there was little further change in early adolescence.

### BMI trajectories are influenced by different early childhood factors

The Early Obesity and Late Weight Gain classes follow clearly distinct and extreme trajectories across all sweeps and represent smaller groups of individuals (3.7% and 3.3%, respectively). The other two classes, Weight Loss and Early Weight Gain, have close starting points at age 3 and then follow less extreme but opposite trajectories during childhood, which represent the majority of individuals (69% and 24%, respectively). Although poverty, ethnic minority, single-parenthood, and low maternal education formed a common core of risk factors for unhealthy weight trajectories, we found different associations with early biological, psychological, and family factors.

### Early socioeconomic context

In our study, children living in economically disadvantaged, ethnic, and single-parent families at 9 months of age were at greater risk of following unhealthy weight trajectories compared to their peers living in white advantaged families. Poverty has been consistently associated with obesity in both children and adults. Indeed, several studies support that early childhood poverty has an enduring association with obesity (35) and that its effect persists despite subsequent improvement in socioeconomic status (36).

However, in our study, this association disappeared after adjusting to other factors like maternal nutritional status and maternal education. In fact, maternal education appears to play a major role in childhood obesity (37). A large prospective study including data from 11 European cohorts concluded that low maternal education substantially increased the risk of early childhood adiposity in all countries (37). Therefore, maternal education might moderate the effect of poverty on obesity.

### Child factors

#### Gestational age, birthweight, breastfeeding, and complementary feeding

In our study, children with high birthweight and who were big for GA were at greater risk for Early Weight Gain and Early Obesity. Interestingly, late prematurity was associated with the Early Weight Gain and Early Obesity trajectories, but extreme to moderate prematurity and being small for GA increased the risk for Late Weight Gain.

Early life context, including pregnancy, has a strong effect on later risk of obesity. Two cross-sectional studies of young adults found that those who had been born before 33 weeks GA had higher adiposity and cardiometabolic risk than those born at term (38,39). Moreover, it has been proposed that both nutrient overabundance and scarcity during pregnancy and infancy lead to a metabolic programming that results in an increased obesity risk throughout the lifespan (40). In fact, in a longitudinal cohort in Rotterdam, individuals with fetal growth restriction followed by infant weight acceleration had higher visceral and liver adiposity than those with normal fetal and infant growth (41).

In our study, being breastfed for more than 2 months lowered the risk of Early Weight Gain and Early Obesity, but not of Late Weight Gain, and early solid food introduction had an opposite effect. These associations were moderated by socioeconomic and biological factors. Breastfeeding has many demonstrated benefits for both mother and child (42). The mechanisms underlying the relationship between breastfeeding and obesity are still debated and include human milk polysaccharides, modulation of gut microbiota, and promotion of sensitive feeding and self-regulation (43,44). The protective effect of exclusive breastfeeding on rapid weight gain is seen mainly in early childhood, but its long-lasting effect has been debated (45). Other authors even argue that this relationship may stem from the more favorable socioeconomic and educational background of breastfeeding mothers (46).

#### Child temperament and self-regulation

In our study, emotional dysregulation in infancy was associated with Early Obesity, even after adjusting for socioeconomic and biological factors.

The effect of infant self-regulation on adolescent obesity has already been demonstrated in a study using the same MCS cohort (25). A growing body of evidence indicates that self-regulation has an important role in eating behavior, with children who show a lower capacity to self-sooth and to self-regulate being at greater risk of using food as a consolation tool and of obesity (47). The ability to regulate emotions begins in infancy and develops in the context of early mother-child reciprocal interactions. Breastfeeding has been shown to promote a mother’s sensitivity and capability of attributing mental states to their babies, and to predict more positive and sensitive behaviors during feeding at 12 months (44).

### Family factors

#### Household routines, parenting beliefs, and parenting activities

In our study, children with consistent sleep and eating routines in early childhood were less likely to follow unhealthy weight trajectories, even after adjusting for socioeconomic and biological factors. The effect of household routines on obesity has already been consistently demonstrated in a study using the MCS and in the USA (25,48).

In our study, more positive parenting beliefs at age 9 months decreased the risk of Late Weight Gain, but this association disappeared after adjusting to other covariates. Although parental engagement in children’s daily activities has been associated with lower obesity risk (49), we found no association between parenting activities and weight trajectories.

#### Discipline practices and child-parent relationship

Children who had a close relationship with their mothers in early childhood were less likely to follow the Late Weight Gain trajectory, and child-parent conflict in early childhood was associated with Early Obesity. There is a strong association between parent-child relationship and unhealthy eating and sedentary behaviors, factors well known to promote weight gain. A recent review highlights that a secure parent-child bond and high parental connectedness are associated with better eating and general health behaviors, while an insecure attachment and difficult parent-child relationship were associated with disordered eating and sedentary behaviors, mediated by temperament and self-regulation (50). Regarding weight status, in a longitudinal study, individuals with poor mother-child relationships in early infancy, assessed by maternal sensitivity and attachment, were at greater odds of obesity during adolescence (51).

In our study, positive parenting showed a small protective association with Early and Late Weight Gain that disappeared after adjusting to other factors. Harsh parenting has been demonstrated to increase the risk of obesity, while the appropriate use of family rules has been associated with a deceleration of obesity risk in adolescence (49,52).

##### Strengths and limitations

This study provided new evidence about the relationship between nutritional status trajectories and child and family factors. Its prospective longitudinal design allowed the exploration of BMI trajectories from early childhood to adolescence, and its large sample size allowed the exploration of multiple important individual and family covariates. To date, this is one of few studies that comprehensively included psychological factors like child temperament, self-regulation, parenting, and child-parent relationship alongside socioeconomic and biological factors.

Also, this study further explored the methodological aspects of investigating BMI trajectories. Anthropometric measures were directly collected according to the best standards, providing reliable and accurate data for subsequent analysis. Furthermore, trajectories were based on a continuous measure of BMI rather than on categorical measures of overweight and obesity. Especially, trajectories were based on BMI Z-scores, considered the gold standard for evaluating anthropometric measures. Moreover, it included 5 repeated measures of BMI over time and explored different GMM specifications.

Nevertheless, this study is not without limitations. We found it challenging to choose the best model. Although there is a common understanding on how model selection should be guided, there is some debate about the best indicators and their cut-offs. Furthermore, we opted for a homoscedastic GMM because the heteroscedastic GMM provided less substantive and poorly defined classes. Nevertheless, we must acknowledge that homoscedasticity might influence our results. Also, non-invariant models, which are more flexible and which have more parameters to estimate, tend to have superior information criteria at the expense of decreased interpretability and entropy, and one might question what would be the use of a better fitting model if it had lost its classification accuracy. Therefore, meaningfulness has a significant role in model selection. Although the entropy of our final model is not ideal, the subsequent regression analysis was adjusted to this classification bias.

Attrition is a major problem in longitudinal studies. GMM uses FIML to estimate the BMI trajectories, including all available data in the analysis, and therefore minimizes the bias effect of attrition.

Child and family covariates showed different percentages of missing values, and a complete case analysis would have substantially reduced the available sample; thus, we needed to impute missing values, which might influence our associations. Multiple imputation was performed according to best practice, and analysis in imputed and non-imputed data yielded concordant results. Although we recategorized covariates according to their distributions, previous research, and, whenever possible, international standards and recommendations, this recategorization might affect the association between BMI trajectories and covariates.

## Conclusion

This study focused on BMI trajectories and looked at their early childhood predictors. In our study, 31% of the children followed an unhealthy weight trajectory that was mainly set by the time they reached early adolescence. Therefore, obesity prevention interventions should target children in early and middle-childhood, particularly those living in disadvantaged families. This study further points out that the lack of routines, low emotional self-regulation, low child-parent closeness, and child-parent conflict are significantly associated with unhealthy BMI trajectories, even after adjusting to other contextual factors; therefore, these are possible areas for family-based, health-promotion interventions. Further studies should focus on how different family and parenting factors interplay and influence weight trajectories, and on possible short- and long-term consequences on health status and well-being, following a developmental perspective.

## Data Availability

The data that support the findings of this study are available through the UK Data Service [https://beta.ukdataservice.ac.uk/datacatalogue/series/series?id=2000031], but restrictions apply to the availability of these data, which were used under Special Licence for the current study. Data are available throught the UK Data Service after approval by the CLS Data Access Committee.

## Abbreviations in alphabetical order

BIC: Schwarz’s Bayesian Information Criterion
BLRT: Bootstrapped likelihood ratio test
BMI: Body mass index
BMIz: Age- and gender-specific BMI Z-scores
FIML: Full information maximum likelihood
GA: Gestational age
GMM: Growth Mixture Modeling
IOTF: International Obesity Task Force
LCGA: Latent Class Growth Analysis
MCS: Millennium Cohort Study
OR: Odds ratio
SD: Standard deviation
WHO: World Health Organization
Z-score: Standard deviation scores

## Acknowledgements

The Millennium Cohort Study (MCS), which began in 2000, is conducted by the Centre for Longitudinal Studies (CLS). It aims to chart the conditions of social, economic and health advantages and disadvantages facing children born at the start of the 21st century. The Principal Investigator and Director of the MCS is Prof. Emla Fitzsimons, UCL Institute of Education, University College London. For details, see https://cls.ucl.ac.uk/cls-studies/millennium-cohort-study/

The authors are grateful to the Centre for Longitudinal Studies (CLS), UCL Institute of Education, for the use of these data and to the UK Data Service for making them available, as well as to all the families who have participated in the MCS. However, neither CLS nor the UK Data Service bear any responsibility for the analysis or interpretation of these data.

## Declarations

### Authors’ contributions

All authors contributed to the study conception and design. Data analysis were performed by CS. The first draft of the manuscript was written by CS and all authors commented on previous versions of the manuscript and revised it critically for important intellectual content. All authors read and approved the final manuscript and agree to be accountable for all aspects of the work in ensuring that questions related to the accuracy or integrity of any part of the work are appropriately investigated and resolved.

## Bibliography

1. NCD Risk Factor Collaboration (NCD-RisC). Worldwide trends in body-mass index, underweight, overweight, and obesity from 1975 to 2016: a pooled analysis of 2416 population-based measurement studies in 1289 million children, adolescents, and adults. Lancet. 2017 Dec 16;390(10113):2627–42.

2. de Onis M, Blössner M, Borghi E. Global prevalence and trends of overweight and obesity among preschool children. Am J Clin Nutr. 2010 Nov;92(5):1257–64.

3. Rodriguez-Ayllon M, Cadenas-Sanchez C, Esteban-Cornejo I, Migueles JH, Mora-Gonzalez J, Henriksson P, et al. Physical fitness and psychological health in overweight/obese children: A cross-sectional study from the ActiveBrains project. J Sci Med Sport. 2017 Oct 4;21(2):179–84.

4. Bacchini D, Licenziati MR, Garrasi A, Corciulo N, Driul D, Tanas R, et al. Bullying and Victimization in Overweight and Obese Outpatient Children and Adolescents: An Italian Multicentric Study. PLoS One. 2015 Nov 25;10(11):e0142715.

5. Patton GC, Coffey C, Carlin JB, Sawyer SM, Williams J, Olsson CA, et al. Overweight and obesity between adolescence and young adulthood: a 10-year prospective cohort study. J Adolesc Health. 2011 Mar;48(3):275–80.

6. Gardner DSL, Hosking J, Metcalf BS, Jeffery AN, Voss LD, Wilkin TJ. Contribution of early weight gain to childhood overweight and metabolic health: a longitudinal study (EarlyBird 36). Pediatrics. 2009 Jan;123(1):e67–73.

7. Hughes AR, Sherriff A, Lawlor DA, Ness AR, Reilly JJ. Timing of excess weight gain in the Avon Longitudinal Study of Parents and Children (ALSPAC). Pediatrics. 2011 Mar;127(3):e730–6.

8. Datar A, Shier V, Sturm R. Changes in body mass during elementary and middle school in a national cohort of kindergarteners. Pediatrics. 2011 Dec;128(6):e1411–7.

9. Birch LL, Davison KK. Family environmental factors influencing the developing behavioral controls of food intake and childhood overweight. Pediatr Clin North Am. 2001 Aug;48(4):893–907.

10. Vollmer RL, Mobley AR. Parenting styles, feeding styles, and their influence on child obesogenic behaviors and body weight. A review. Appetite. 2013 Dec;71:232–41.

11. Sleddens EFC, Gerards SMPL, Thijs C, de Vries NK, Kremers SPJ. General parenting, childhood overweight and obesity-inducing behaviors: a review. Int J Pediatr Obes. 2011 Jun 9;6(2-2):e12–27.

12. Halliday JA, Palma CL, Mellor D, Green J, Renzaho AMN. The relationship between family functioning and child and adolescent overweight and obesity: a systematic review. Int J Obes. 2014 Apr;38(4):480–93.

13. Mattsson M, Maher GM, Boland F, Fitzgerald AP, Murray DM, Biesma R. Group-based trajectory modelling for BMI trajectories in childhood: A systematic review. Obes Rev. 2019 Jul;20(7):998–1015.

14. Muthén B, Muthén LK. Integrating person-centered and variable-centered analyses: growth mixture modeling with latent trajectory classes. Alcohol Clin Exp Res. 2000 Jun;24(6):882–91.

15. Mei Z, Grummer-Strawn LM. Standard deviation of anthropometric Z-scores as a data quality assessment tool using the 2006 WHO growth standards: a cross country analysis. Bull World Health Organ. 2007 Jun;85(6):441–8.

16. Connelly R, Platt L. Cohort profile: UK Millennium Cohort Study (MCS). Int J Epidemiol. 2014 Dec;43(6):1719–25.

17. Department of Nutrition for Health and Development at the World Health Organization (WHO). WHO Anthro Survey Analyser: Software for analysing survey anthropometric data for children under 5 years of age. [Internet]. Geneva: World Health Organization; 2018 [cited 2019 Oct 25]. Available from: https://whonutrition.shinyapps.io/anthro/

18. WHO. WHO AnthroPlus for personal computers Manual: Software for assessing growth of the world’s’ ‘ children and adolescents. [Internet]. Geneva: WHO; 2019 [cited 2019 Oct 25]. Available from: https://www.who.int/growthref/tools/en/

19. March of Dimes, PMNCH, Save the children, WHO. Born Too Soon: The Global action report on preterm Birth [Internet]. Geneva: CP Howson, MV Kinney, Jelawn. World Health Organization.; 2012 [cited 2019 Oct 25]. Available from: https://www.who.int/maternal_child_adolescent/documents/born_too_soon/en/

20. World Health Organization. ICD-10?: international statistical classification of diseases and related health problems?: tenth revision. World Health Organization; 2004.

21. Villar J, Cheikh Ismail L, Victora CG, Ohuma EO, Bertino E, Altman DG, et al. International standards for newborn weight, length, and head circumference by gestational age and sex: the Newborn Cross-Sectional Study of the INTERGROWTH-21st Project. Lancet. 2014 Sep 6;384(9946):857–68.

22. Carling SJ, Demment MM, Kjolhede CL, Olson CM. Breastfeeding duration and weight gain trajectory in infancy. Pediatrics. 2015 Jan;135(1):111–9.

23. Fewtrell M, Bronsky J, Campoy C, Domellöf M, Embleton N, Fidler Mis N, et al. Complementary feeding: A position paper by the european society for paediatric gastroenterology, hepatology, and nutrition (ESPGHAN) committee on nutrition. J Pediatr Gastroenterol Nutr. 2017;64(1):119–32.

24. Hernández-Alava M, Popli G. Children’s development and parental input: evidence from the UK millennium cohort study. Demography. 2017;54(2):485–511.

25. Anderson SE, Sacker A, Whitaker RC, Kelly Y. Self-regulation and household routines at age three and obesity at age eleven: longitudinal analysis of the UK Millennium Cohort Study. Int J Obes. 2017 Apr 24;41(10):1459–66.

26. Kroll ME, Carson C, Redshaw M, Quigley MA. Early father involvement and subsequent child behaviour at ages 3, 5 and 7 years: prospective analysis of the UK millennium cohort study. PLoS One. 2016 Sep 21;11(9):e0162339.

27. Rajyaguru P, Moran P, Cordero M, Pearson R. Disciplinary parenting practice and child mental health: evidence from the UK millennium cohort study. J Am Acad Child Adolesc Psychiatry. 2019;58(1):108–116.e2.

28. Nylund KL, Asparouhov T, Muthén BO. Deciding on the Number of Classes in Latent Class Analysis and Growth Mixture Modeling: A Monte Carlo Simulation Study. Structural Equation Modeling: A Multidisciplinary Journal. 2007 Oct 23;14(4):535–69.

29. Todo N, Usami S. Fitting unstructured finite mixture models in longitudinal design: A recommendation for model selection and estimation of the number of classes. Structural Equation Modeling: A Multidisciplinary Journal. 2016 Sep 2;23(5):695–712.

30. Celeux G, Soromenho G. An entropy criterion for assessing the number of clusters in a mixture model. J of Classification. 1996 Sep;13(2):195–212.

31. Vermunt JK. Latent Class Modeling with Covariates: Two Improved Three-Step Approaches. Political Analysis. 2010;18(04):450–69.

32. Lang KM, Little TD. Principled missing data treatments. Prev Sci. 2016 Apr 4;19(3):1–11.

33. Kelly Y, Patalay P, Montgomery S, Sacker A. BMI Development and Early Adolescent Psychosocial Well-Being: UK Millennium Cohort Study. Pediatrics. 2016 Nov 11;138(6).

34. Stuart B, Panico L. Early-childhood BMI trajectories: evidence from a prospective, nationally representative British cohort study. Nutr Diabetes. 2016 Mar 7;6:e198.

35. Lee H, Andrew M, Gebremariam A, Lumeng JC, Lee JM. Longitudinal associations between poverty and obesity from birth through adolescence. Am J Public Health. 2014 May;104(5):e70–6.

36. Li M, Mustillo S, Anderson J. Childhood poverty dynamics and adulthood overweight/obesity: Unpacking the black box of childhood. Soc Sci Res. 2018 Nov;76:92–104.

37. Ruiz M, Goldblatt P, Morrison J, Porta D, Forastiere F, Hryhorczuk D, et al. Impact of low maternal education on early childhood overweight and obesity in europe. Paediatr Perinat Epidemiol. 2016 May;30(3):274–84.

38. Thomas EL, Parkinson JR, Hyde MJ, Yap IKS, Holmes E, Doré CJ, et al. Aberrant adiposity and ectopic lipid deposition characterize the adult phenotype of the preterm infant. Pediatr Res. 2011 Nov;70(5):507–12.

39. Sipola-Leppänen M, Vääräsmäki M, Tikanmäki M, Matinolli H-M, Miettola S, Hovi P, et al. Cardiometabolic risk factors in young adults who were born preterm. Am J Epidemiol. 2015 Jun 1;181(11):861–73.

40. Koletzko B, Brands B, Grote V, Kirchberg FF, Prell C, Rzehak P, et al. Long-Term Health Impact of Early Nutrition: The Power of Programming. Ann Nutr Metab. 2017 Jul 6;70(3):161–9.

41. Vogelezang S, Santos S, Toemen L, Oei EHG, Felix JF, Jaddoe VWV. Associations of fetal and infant weight change with general, visceral, and organ adiposity at school age. JAMA Netw Open. 2019 Apr 5;2(4):e192843.

42. Victora CG, Bahl R, Barros AJD, França GVA, Horton S, Krasevec J, et al. Breastfeeding in the 21st century: epidemiology, mechanisms, and lifelong effect. Lancet. 2016 Jan 30;387(10017):475–90.

43. Alderete TL, Autran C, Brekke BE, Knight R, Bode L, Goran MI, et al. Associations between human milk oligosaccharides and infant body composition in the first 6 mo of life. Am J Clin Nutr. 2015 Dec;102(6):1381–8.

44. Farrow C, Blissett J. Maternal mind-mindedness during infancy, general parenting sensitivity and observed child feeding behavior: a longitudinal study. Attach Hum Dev. 2014 Mar 31;16(3):230–41.

45. Liu JX, Xu X, Liu JH, Hardin JW, Li R. Association of maternal gestational weight gain with their offspring’s anthropometric outcomes at late infancy and 6 years old: Mediating roles of birth weight and breastfeeding duration. Int J Obes. 2017 Aug 4;42(1):8–14.

46. Coduti N, Gregoire M, Sowa D, Diakakis GM, Chen Y. Characteristics of exclusively breastfeeding mothers. Top Clin Nutr. 2015;30(2):174–83.

47. Graziano PA, Kelleher R, Calkins SD, Keane SP, Brien MO. Predicting weight outcomes in preadolescence: the role of toddlers’ self-regulation skills and the temperament dimension of pleasure. Int J Obes. 2013 Jul;37(7):937–42.

48. Anderson SE, Whitaker RC. Household routines and obesity in US preschool-aged children. Pediatrics. 2010 Mar;125(3):420–8.

49. Huang DYC, Lanza HI, Anglin MD. Trajectory of adolescent obesity: exploring the impact of prenatal to childhood experiences. J Child Fam Stud. 2014 Aug 1;23(6):1090–101.

50. Blewitt C, Bergmeier H, Macdonald JA, Olsson CA, Skouteris H. Associations between parent-child relationship quality and obesogenic risk in adolescence: a systematic review of recent literature. Obes Rev. 2016 Jul;17(7):612–22.

51. Anderson SE, Gooze RA, Lemeshow S, Whitaker RC. Quality of early maternal-child relationship and risk of adolescent obesity. Pediatrics. 2012 Jan;129(1):132–40.

52. Lohman BJ, Gillette MT, Neppl TK. Harsh parenting and food insecurity in adolescence: the association with emerging adult obesity. J Adolesc Health. 2016 Jul;59(1):123–7.

## Data references

1. University of London, Institute of Education, Centre for Longitudinal Studies. (2017). Millennium Cohort Study: Longitudinal Family File, 2001-2015. [data collection]. 2nd Edition. UK Data Service. SN:8172, http://doi.org/10.5255/UKDA-SN-8172-2

2. University of London, Institute of Education, Centre for Longitudinal Studies. (2017). Millennium Cohort Study: First Survey, 2001-2003. [data collection]. 12th Edition. UK Data Service. SN: 4683, http://doi.org/10.5255/UKDA-SN-4683-4

3. University of London, Institute of Education, Centre for Longitudinal Studies. (2017). Millennium Cohort Study: Second Survey, 2003-2005. [data collection]. 9th Edition. UK Data Service. SN: 5350, http://doi.org/10.5255/UKDA-SN-5350-4

4. University of London, Institute of Education, Centre for Longitudinal Studies. (2017). Millennium Cohort Study: Third Survey, 2006. [data collection]. 7th Edition. UK Data Service. SN: 5795, http://doi.org/10.5255/UKDA-SN-5795-4

5. University of London, Institute of Education, Centre for Longitudinal Studies. (2017). Millennium Cohort Study: Fourth Survey, 2008. [data collection]. 7th Edition. UK Data Service. SN: 6411, http://doi.org/10.5255/UKDA-SN-6411-7

6. University of London, Institute of Education, Centre for Longitudinal Studies. (2017). Millennium Cohort Study: Fifth Survey, 2012. [data collection]. 4th Edition. UK Data Service. SN: 7464, http://doi.org/10.5255/UKDA-SN-7464-4

7. University of London, Institute of Education, Centre for Longitudinal Studies. (2019). Millennium Cohort Study: Sixth Survey, 2015. [data collection]. 4th Edition. UK Data Service. SN: 8156, http://doi.org/10.5255/UKDA-SN-8156-4

